# Outbreak of severe community-acquired bacterial infections from *Streptococcus pyogenes, Streptococcus pneumoniae, Neisseria meningitidis*, and *Haemophilus influenzae* among children in North Rhine-Westphalia (Germany), October to December 2022

**DOI:** 10.1101/2023.09.14.23295531

**Authors:** Sarah C. Goretzki, Mark van der Linden, Andreas Itzek, Tom Hühne, Roland O. Adelmann, Firas Ala Eldin, Mohamed Albarouni, Jan-Claudius Becker, Michael A. Berghäuser, Thomas Boesing, Michael Boeswald, Milian Brasche, Francisco Brevis, Rokya Camara, Clara Deibert, Frank Dohle, Jörg Dolgner, Jan Dziobaka, Frank Eifinger, Natalie Elting, Matthias Endmann, Guido Engelmann, Holger Frenzke, Monika Gappa, Bahman Gharavi, Christine Goletz, Eva Hahn, Yvonne Heidenreich, Konrad Heimann, Kai O. Hensel, Hans-Georg Hoffmann, Marc Hoppenz, Gerd Horneff, Helene Klassen, Cordula Körner-Rettberg, Alfred Längler, Pascal Lenz, Klaus Lohmeier, Andreas Müller, Frank Niemann, Michael Paulussen, Falk Pentek, Ruy Perez, Markus Pingel, Philip Repges, Tobias Rothoeft, Jochen Rübo, Herbert Schade, Robert Schmitz, Peter Schonhoff, Jan N. Schwade, Tobias Schwarz, Peter Seiffert, Georg Selzer, Uwe Spille, Carsten Thiel, Ansgar Thimm, Bartholomäus Urgatz, Alijda van den Heuvel, Tan van Hop, Verena Giesen, Stefan Wirth, Thomas Wollbrink, Daniel Wüller, Ursula Felderhoff-Müser, Christian Dohna-Schwake, Thiên-Trí Lâm, Heike Claus, N. Bruns

**Author notes:** **Address correspondence to**: Nora Bruns, Department of Pediatrics I (Neonatology, Pediatric Intensive Care, Pediatric Neurology, and Pediatric Infectious Diseases), University Hospital, Essen, University of Duisburg-Essen, Holsterhauserstr. 172, 40547 Essen, Germany, [ ]. **Funding/Support:** This research received no external funding. The surveillance of invasive streptococcal infections in Germany at the German National Reference Laboratory for Streptococci was funded by the Robert Koch Institute, Berlin, Germany (funding number: 1369-235). **Role of Funder/Sponsor (if any):** The funder had no role in the design and conduct of the study. **Clinical Trial Registration (if any)** n/a.

## Abstract

**Background:** In late 2022, a surge of severe bacterial infections caused by *S. pyogenes* was reported in several European countries, including Germany. This study assessed disease burden and severity of hospitalizations for community-acquired bacterial infections with *S. pyogenes, S. pneumoniae, N. meningitidis*, and *H. influenzae* among children in North Rhine-Westphalia (NRW), Germany, during the last quarter of 2022 compared to long-term incidences.

**Methods:** Hospital cases due to bacterial infections between October and December 2022 were collected from 59/62 (95 %) children’s hospitals in NRW and combined with surveillance data (2016 - 2023) from the national reference laboratories for streptococci, *N. meningitidis*, and *H. influenzae*. Total cases in NRW and incidence rates from January 2016 to March 2023 were estimated by capture-recapture analyses. Expected annual deaths from the studied pathogens were calculated from national death cause statistics.

**Results:** Between October and December 2022, 153 cases with high overall disease severity were reported with pneumonia being most common (59 %, n = 91). Incidence rates of bacterial infections declined at the beginning of the COVID-19 pandemic. In late 2022 and early 2023 a massive surge to levels unprecedented since 2016 was observed, mainly driven by *S. pyogenes* and *S. pneumoniae*. Observed deaths during the study period exceeded the expected number for the entire year in NRW by far (7 vs. 0.9).

**Discussion:** The unprecedented peak of bacterial infections in late 2022 and early 2023 was caused by various mechanisms intertwined that require close surveillance and improved precautionary measures for future outbreaks.

## Introduction

Since the outbreak of the COVID-19 pandemic in 2020, children’s and adolescents’ health has been challenged in numerous ways. Among others, a decline of bacterial infections and distorted periodicity of seasonal infection waves were observed across the world and affected all age groups.^1–5^ An unforeseen result of reduced incidence of infections is the lack of natural immunity especially in the youngest.

Beginning in late fall 2022, rising numbers of severe bacterial infections caused by group A streptococci were reported from Spain, France, Great Britain, and other countries.^6–9^ At the same time in Germany, a massive wave of acute viral and bacterial infections caused a near-collapse of the pediatric health care system including out- and inpatient sectors and - most dramatically - overwhelmed pediatric intensive care capacities.^10–12^ Pediatric health care providers in Germany claimed that this was an unprecedented public health emergency.

This study was designed to assess the burden of hospitalizations due to community-acquired bacterial infections by *Streptococcus pyogenes* (*S. pyogenes*), *Streptococcus pneumoniae* (*S. pneumoniae*), *Neisseria meningitidis* (*N. meningitidis*), and *Haemophilus influenzae* (*H. influenzae*) in the last quarter of 2022 compared to seasonally expected infection waves since 2016; to identify driving pathogens, and describe disease severity and outcomes. Accordingly, we combined five data sources for comprehensive capture-recapture analyses: 1) A multicentre study (MC) conducted in 95 % of children’s hospitals in the most populated state of Germany (North Rhine-Westphalia [NRW]) in the last quarter of 2022; 2) long-term surveillance data from national reference laboratories for a) streptococci, b) *N. meningitidis*, and c) *Haemophilus influenzae*; as well as 3) publicly accessible population and death cause statistics provided by the German Federal Statistical Office (FSO, Statistisches Bundesamt, www.destatis.de).

## Methods

### Study design

This is a retrospective multicentre cohort study to assess the burden of community-acquired severe bacterial infections in children and adolescents in NRW/Germany (NRW) between October 1^st^ and December 31^st^ of 2022. The results were combined with four data sources to deduce long-term trends.

First, clinical data were collected from 60 of 63 (95 %) children’s hospitals in NRW. Second, surveillance data from the national reference laboratories (NRL) for streptococci, *N. meningitidis*, and *H. influenzae* were matched with clinical cases from the same period to conduct capture-recapture (CRC) analyses for each pathogen. Third, monthly NRL-reported incidence numbers between January 2016 and March 2023 were combined with population statistics published by the FSO to calculate incidence rates and ratios. Fourth, death statistics by the FSO were accessed to calculate the expected number of deaths by bacterial infections per year and compare them with the numbers observed in the MC study.

### Multicentre study

#### Study population

Children aged > 27 days and < 18 years admitted to a children’s hospital in NRW during the study period due to a community-acquired infection with *S. pyogenes*, *S. pneumoniae*, *N. meningitidis*, or *H. influenzae* were eligible. Children with catheter-associated infections from permanently indwelling catheters and cases of nosocomial infections were excluded.

#### Clinical data collection

Executives of all children’s hospitals in NRW were invited to participate. Eligible patients were identified via positive test results for the bacterial pathogen of interest by the local microbiology departments or derived from diagnose related group codes (International Classification of Diseases, 10^th^ revision, German modification). Eligibility was confirmed via retrospective chart review by the local investigators. Anonymized clinical data were collected via web-based case report forms (eCRF) hosted at www.limesurvey.org.

#### Handling of duplicates

Duplicate reports in the dataset originating from inter-centre referrals were identified by discharge date information and destination of referral in the eCRF. The total duration of hospital stay in both hospitals was updated along with clinical information, and the duplicate case removed.

Children’s hospital capacities of North Rhine-Westphalia:

Each participating hospital reported the number of non-surgical beds on pediatric wards (including neonatal and pediatric intensive care units) during the study period in the eCRFs. The capacities of the three non-participating centres were obtained by personal inquiry to the attending office of the respective children’s hospital.

### National surveillance of invasive bacterial infections

#### Streptococci

Reporting of invasive *S. pneumoniae* infections is mandatory in Germany. However, surveillance of invasive streptococcal disease by the German national reference laboratory for streptococci (NRLS) is based on voluntary reporting by primary diagnostic laboratories, which are quested to send all streptococcal isolates from invasive infections (isolate sampled from a normally sterile anatomical site) to the NRLS for detailed analyses. This includes mainly microbiological- (hemolysis- and colony-size assessment, optochin sensitivity as well as bile solubility), biochemical- (leucine-aminopeptidase- and pyrrolidonylarylamidase-test), serological- (Neufeld-Quellung reaction) and molecularbiological methods (*16S rRNA* sequencing, *emm*-typing).

#### N. meningitidis and H. influenzae

Mandatory reporting is required for cases of meningococcal meningitis and meningococcal septicaemia. Additionally, the detection of *N. meningitidis* in blood, cerebrospinal fluid, petechiae, or other sterile sites, as well as the detection of *H. influenzae* in blood or cerebrospinal fluid, must be reported. While it is voluntary to submit bacterial isolates or clinical specimens to the National Reference Laboratory for *N. meningitidis* and *H. influenzae* (NRLMHi), up to 90% of the reported cases are covered by the submissions to NRLMHi. Following species confirmation, meningococcal serogroups and *H. influenzae* serotypes are determined through slide agglutination using specific commercial antibodies.

### Population statistics of NRW

The end of year-populations of NRW and Germany from 2015 through 2022 were accessed at the FSO homepage (www.destatis.de). This information was used to calculate midyear populations as well as inhabitant percentage of children < 18 years of age.

National death cause statistics:

Nationwide death cause statistics were accessed at the FSO homepage and the yearly numbers of deaths from streptococci, *N. meningitidis*, and *H. influenzae* infections were extracted for 2016 to 2021 based on the diagnose code of the international classification of diseases 10 German modification (ICD-10-GM) (Table 1). Because age grouping available at the FSO does not comply with the cut-off age for treatment in pediatric departments, expected deaths were calculated only for children < 15 years of age. The number of nationwide expected deaths per year was extrapolated based on the proportion of the pediatric population of NRW (22 %) compared to the total pediatric population of Germany.

**Table 1:**
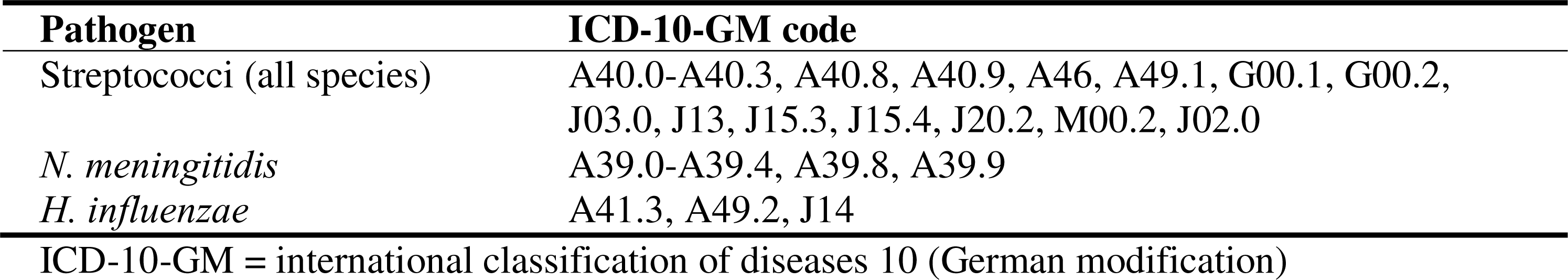
International classification of diseases codes used to calculate the number of expected deaths from bacterial infections in children in Germany.

### Linkage of clinical and surveillance data

Clinical MC and NRL surveillance data of *S. pyogenes*, *S. pneumoniae, N. meningitidis and H. influenzae w*ere matched by patient’s age, sex, sample collection site and date, identified pathogen and serotype if applicable. Direct record linkage by name or date of birth was inapplicable due to anonymization and prohibited by general data protection regulations.

### Statistical analyses

Continuous variables are presented as means with 95 % confidence intervals (CI) if evenly distributed and as median with interquartile range (IQR) if skewed. Discrete variables are presented as counts and relative frequencies. Descriptive statistics were stratified by pathogen. The dataset of the MC study contained no missing data.

Capture-recapture analyses (CRC) were conducted using a generalized log-linear model with Poisson distribution to estimate the number of non-captured cases with 95 % CIs and calculated the estimated number of total cases. Stratification to account for age-, sex- or centre-specific probabilities could not be performed due to the overall low case numbers.

Monthly incidence rates per 100.000 child years (CY) were derived from total case numbers estimated by CRC analyses assuming a consistent reporting rate over time. Monthly incidence rate ratios (IRR) were calculated for the period from January 2020 to March 2023, with the corresponding months of 2016-2019 as references. Besides pathogen-specific incidence rates and IRRs, cumulative monthly incidence rates and IRRs for all four pathogens were calculated to assess the overall burden. 95 % CIs for rates and rate ratios were calculated based on the Poisson distribution.

SAS Enterprise Guide 8.3 (SAS Institute Inc., Cary, NC, USA) was used to perform statistical analyses and produce figures.

### Ethics approval

The study was approved by the Ethics Committee of the Medical Faculty of the University of Duisburg-Essen (22-11045-BO).

## Results

Fifty-nine of 62 (95 %) children’s hospitals in NRW participated in the MC study, comprising 4066 of 4323 (94 %) self-reported pediatric hospital beds during the study period. At the end of 2022, 22 % of Germany’s pediatric population lived in NRW. After subtracting the 6 % capacities from non-participating children’s hospitals, the participating centers supplied medical care for approximately 20.7 % of the German pediatric population.

A total of 153 cases were reported with a median patient age of 4 years (IQR 1 – 7) and 56 % (n = 86) males (Table 2). Fifty-eight (38 %) of the reported cases were associated with *S. pyogenes*, 62 (41 %) with *S. pneumoniae*, 2 (1 %) with *N. meningitidis*, 19 (12 %) with *H. influenzae*, and 12 (8 %) with other streptococcal species. The most frequent presentation was pneumonia (n = 91, 59 %), followed by sepsis/systemic inflammatory response syndrome/toxic shock syndrome (n = 28, 18 %), skin or soft tissue infections (n = 22, 14 %), and ear-nose-throat infections without cerebral invasion (n = 22, 14 %) (Table 2). Viral co-infections were observed in 71 (46%) cases, with variations depending on the bacterial pathogen (Table 2). Intensive care unit admissions were frequent (n = 90, 59%), as was mechanical ventilation (n = 67, 44 %), surgical source control (n = 41, 27 %), and use of vasopressors or inotropes (n = 26, 17 %) (Table 2). Substantial variation between pathogens was observed regarding clinical presentation and performed supportive therapies (Figure 1). Eight children died (5 %), and five patients (3 %) were discharged to a rehabilitation or care facility. Nonetheless, overall functional neurological outcome at hospital discharge in survivors was good (median pediatric cerebral performance category (PCPC) = 1, IQR 1 – 2).

**Table 2:** Clinical characteristics of hospital admissions due to bacterial infections reported in a multicentre study in children’s hospitals in North Rhine-Westphalia (Germany) between October 1^st^ and December 31^st^, 2022.

|  | Total | *S. pyogenes* | *S. pneumoniae* | *H. influenzae* | *N. meningitidis* | *S. spp.\** |
| --- | --- | --- | --- | --- | --- | --- |
|  | N = 153<br>(100%) | n = 58<br>(38 %) | n = 62<br>(41 %) | n = 19<br>(12 %) | n = 2<br>(1 %) | n = 12<br>(8 %) |
| Male, *n* (%) | 86 (56%) | 39 (67 %) | 30 (48 %) | 10 (53 %) | 1 (50 %) | 6 (50 %) |
| Age in years, *median (IQR)* | 4 (1 – 7) | 4 (2 – 7) | 2.5 (1 – 5) | 3 (0 – 6) | 9.5 (2 - 17) | 3.5 (0.5 – 8.5) |
| Viral co-infection, *n* (%) | 71 (46 %) | 18 (31 %) | 37 (60 %) | 14 (74 %) | 0 (0 %) | 2 (17 %) |
| RSV, *n* (%) | 21 (14 %) | 2 (3 %) | 10 (16 %) | 8 (41 %) | 0 (0 %) | 1 (8 %) |
| Influenza A/B, *n* (%) | 29 (19 %) | 12 (21 %) | 14 (23 %) | 3 (16 %) | 0 (0 %) | 0 (0 %) |
| SARS-CoV-2, *n* (%) | 3 (2 %) | 1 (2 %) | 2 (3 %) | 0 (0 %) | 0 (0 %) | 0 (0 %) |
| Other, *n* (%) | 24 (16 %) | 4 (7 %) | 17 (27 %) | 3 (16 %) | 0 (0 %) | 1 (8 %) |
| Bacterial co-infection with *H. influenzae*, *n* (%) | 12 (8 %) | 1 (2 %) | 10 (16 %) | - | 0 (0 %) | 1 (8 %) |
| Clinical presentation |  |  |  |  |  |  |
| Sepsis, SIRS, Toxic Shock, *n* (%) | 28 (18 %) | 18 (31 %) | 1 (2 %) | 1 (5 %) | 1 (50 %) | 7 (58 %) |
| ENT infections w/o cerebral invasion, *n* (%) | 22 (14 %) | 16 (28 %) | 3 (5 %) | 2 (11 %) | 0 (0 %) | 3 (25 %) |
| ENT infections w/ cerebral invasion, *n* (%) | 15 (5 %) | 9 (2 %) | 6 (10 %) | 0 (0 %) | 0 (0 %) | 0 (0 %) |
| CNS infection, *n* (%) | 15 (10 %) | 5 (9 %) | 4 (7 %) | 0 (0 %) | 2 (100 %) | 4 (33 %) |
| Pneumonia, *n* (%) | 91 (59 %) | 22 (38 %) | 51 (82 %) | 15 (79 %) | 0 (0 %) | 3 (25 %) |
| Skin and soft tissue infections, *n* (%) | 22 (14 %) | 17 (29 %) | 4 (7 %) | 0 (0 %) | 0 (0 %) | 1 (8 %) |
| Other infections, *n* (%) | 5 (3 %) | 2 (3 %) | 0 (0 %) | 1 (5 %) | 0 (0 %) | 3 (25 %) |
| Treatment |  |  |  |  |  |  |
| Resuscitation, *n* (%) | 6 (4 %) | 5 (9 %) | 0 (0 %) | 1 (5 %) | 0 (0 %) | 0 (0 %) |
| PICU admission, *n* (%) | 90 (59 %) | 29 (50 %) | 42 (68 %) | 12 (63 %) | 2 (100 %) | 4 (33 %) |
| Invasive ventilation, *n* (%) | 67 (44 %) | 23 (40 %) | 28 (45 %) | 12 (63 %) | 1 (50 %) | 3 (25 %) |
| Administration of vasopressors/inotropes, *n* (%) | 26 (17 %) | 14 (24 %) | 7 (11 %) | 5 (26 %) | 0 (0 %) | 0 (0 %) |
| Extracorporeal membrane oxygenation, *n* (%) | 1 (1 %) | 1 (2 %) | 0 (0 %) | 0 (0 %) | 0 (0 %) | 0 (0 %) |
| Surgical source control, *n* (%) | 41 (27 %) | 22 (38 %) | 14 (23 %) | 0 (0 %) | 0 (0 %) | 7 (58 %) |
| Length of hospital stay hospital, *median (IQR)* | 10 (5 – 19) | 8 (4 – 19) | 11 (5 – 19) | 5 (4 – 6) | 6 (1 - 11) | 13 (7.5 – 25.5) |
| Discharge to rehabilitation/care facility, *n* (%) | 5 (3 %) | 2 (3 %) | 2 (3 %) | 0 (0%) | 0 (0%) | 1 (8 %) |
| PCPC at hospital discharge, *median (IQR)* | 1 (1 – 2) | 1 (1 – 2) | 1 (1 – 2) | 1 (1 – 2) | 1 (1 – 1) | 1 (1 – 2) |
| Deceased, *n* (%) | 8 (5 %) | 5 (9 %) | 2 (3 %) | 1 (5 %) | 0 (0 %) | 0 (0 %) |

\*Streptococcal species other than *S. pyogenes* and *S. pneumoniae*, IQR = interquartile range; ENT = ears nose and throat; CNS = central nervous system; SIRS = systemic inflammatory response syndrome, PICU = pediatric intensive care unit; PCPC = Pediatric Cerebral Performance Category

**Figure 1.**
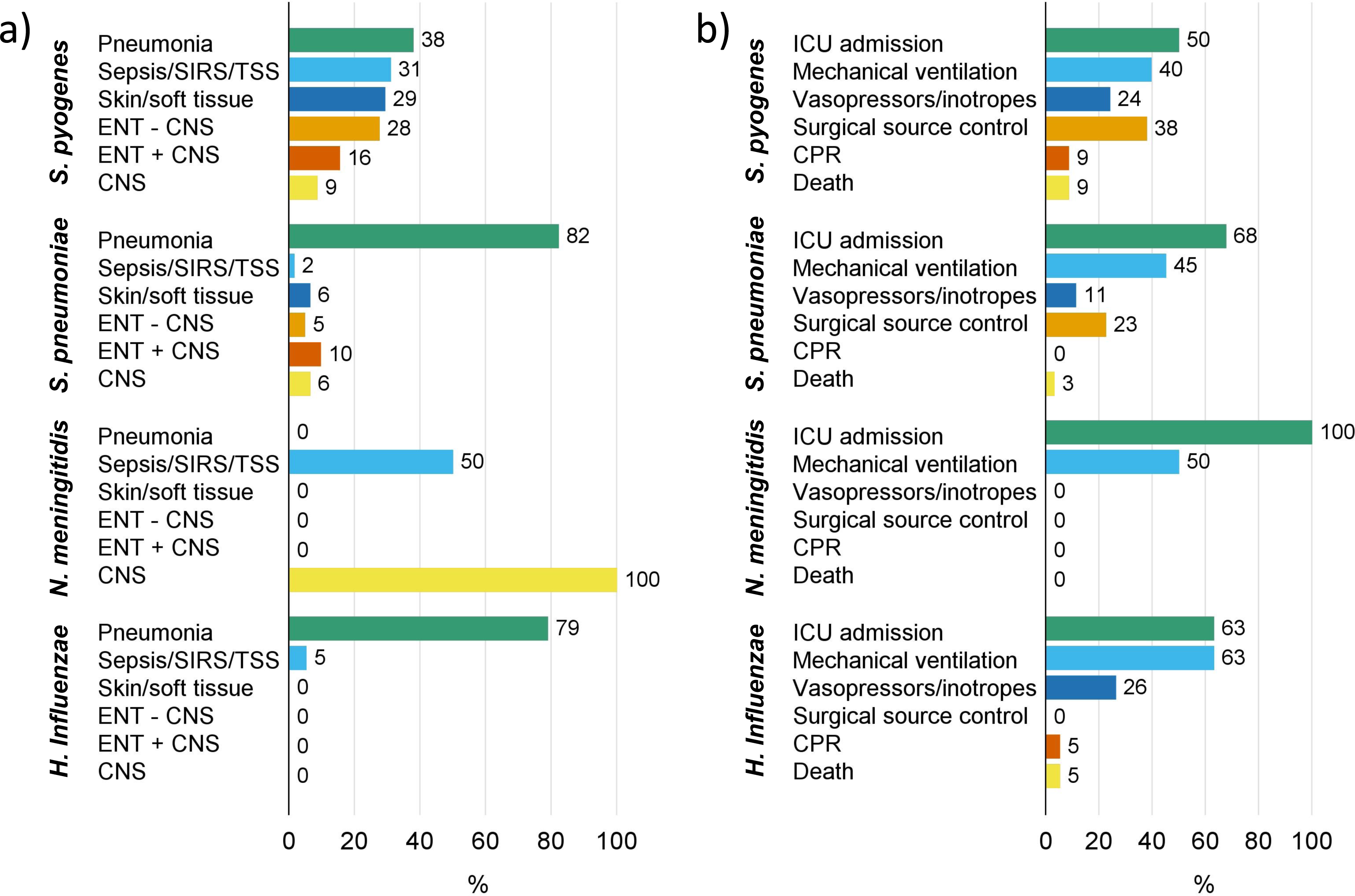
Hospital-treated community-acquired bacterial infections in children in North Rhine-Westphalia in the last quarter of 2022. a) Clinical presentation b) Treatment and outcomes SIRS = systemic inflammatory response syndrome; TSS = toxic shock syndrome, ENT = ear-nose-throat; CNS = central nervous system; ICU = intensive care unit; CPR = cardiopulmonary resuscitation

Capture-recapture analyses showed different degrees of overlap between the conducted MC study and the NRL surveillance data. For *N. meningitidis*, complete capture by the NRL was achieved, but the total case number (n = 6) was comparatively low, especially when compared to case estimates of *S. pneumoniae* (n_estimated_ = 232), *S. pyogenes* (n_estimated_ = 214), and *H. influenzae* (n_estimated_ = 95) (Table 3).

**Table 3:**
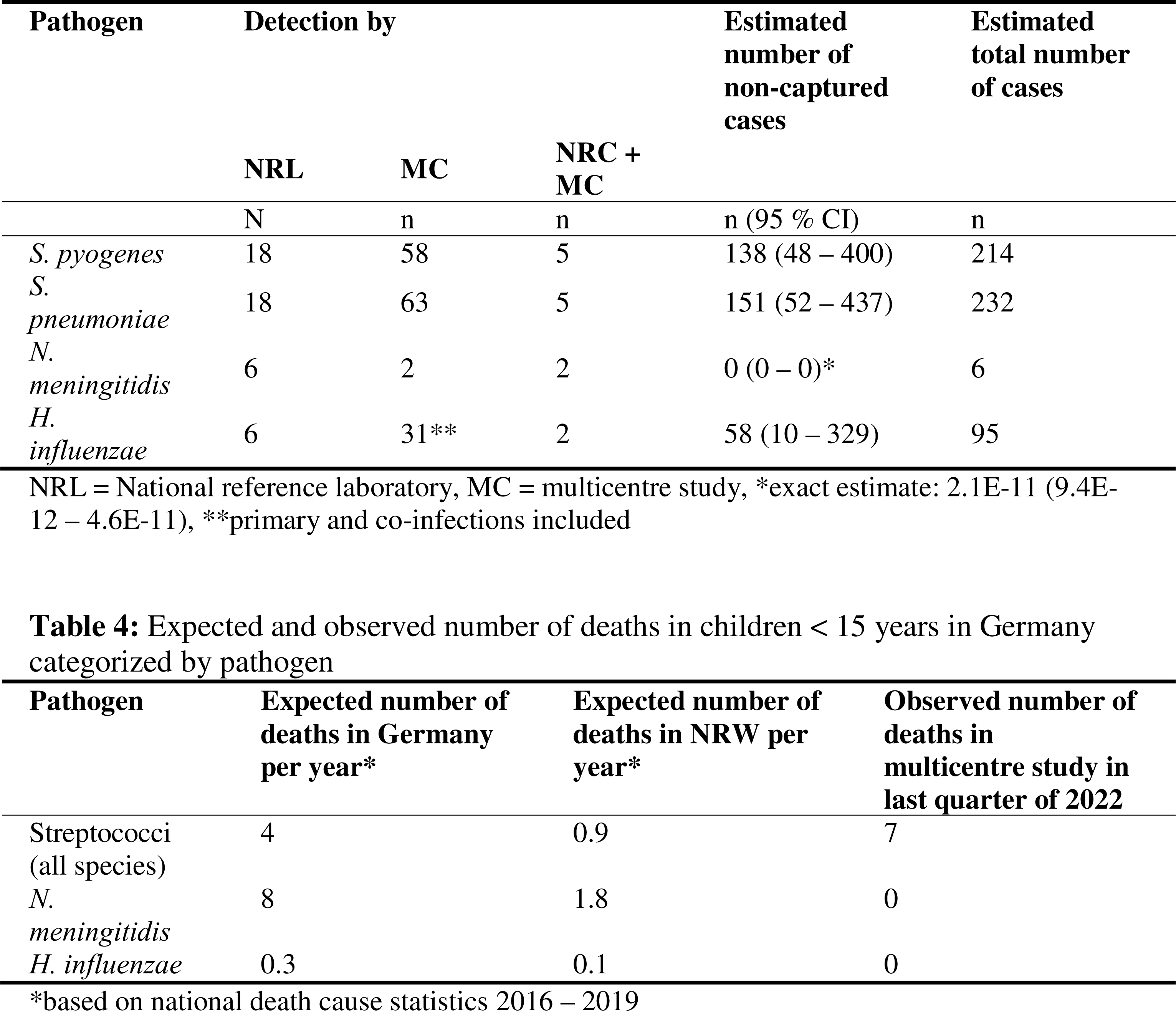
National surveillance and multicentre study detection of pediatric severe and invasive bacterial infections between October 1^st^ and December 31^st^ of 2022 in North-Rhine Westphalia (Germany)

With onset of the COVID-19 pandemic, monthly *S. pyogenes* case numbers reported to the NRLS, monthly incidence rates and IRRs dropped, remained low until September 2022 and peaked in December 2022 (IRR 18.3 (95 % CI 6.3 – 88.8)) as well as March 2023 (IRR 14.7 (5.7 – 54.0)) (Figure 2a and S1a). *S. pneumoniae* also showed a decline during pandemic years but seasonal variations remained present throughout the three-year period, with a remarkable peak in December 2022 (IRR 2.7 (1.7 – 4.7)) and February 2023 (IRR 3.2 (2.0 – 5.5)) (Figure 2b and S1b). For *N. meningitidis*, overall case numbers were fairly low and no clear trend was observed throughout the observation period except for an increase in March 2023 (IRR 1.7 (1.0 – 3.1)) (Figure 2c and S1c). *H. influenzae*, on the other hand, remained mainly within the seasonally expected ranges except for November 2022 (IRR 7.8 (2.2 – 64.8)) and February 2023 (IRR 2.9 (1.1 – 10.8)) (Figure 2d and S1c). Overall, incidence rates and IRRs exhibited a decline at the onset of the pandemic, followed by a pathogen-specific degree of recovery, reaching or surpassing pre-pandemic levels (Figure S1 and supplementary table). Cumulative monthly incidence rates and IRRs declined at the beginning of the pandemic, showed small peaks during the course of the pandemic, and peaked in December 2022 and February 2023 (Figure 3 and supplementary table).

**Figure 2.**
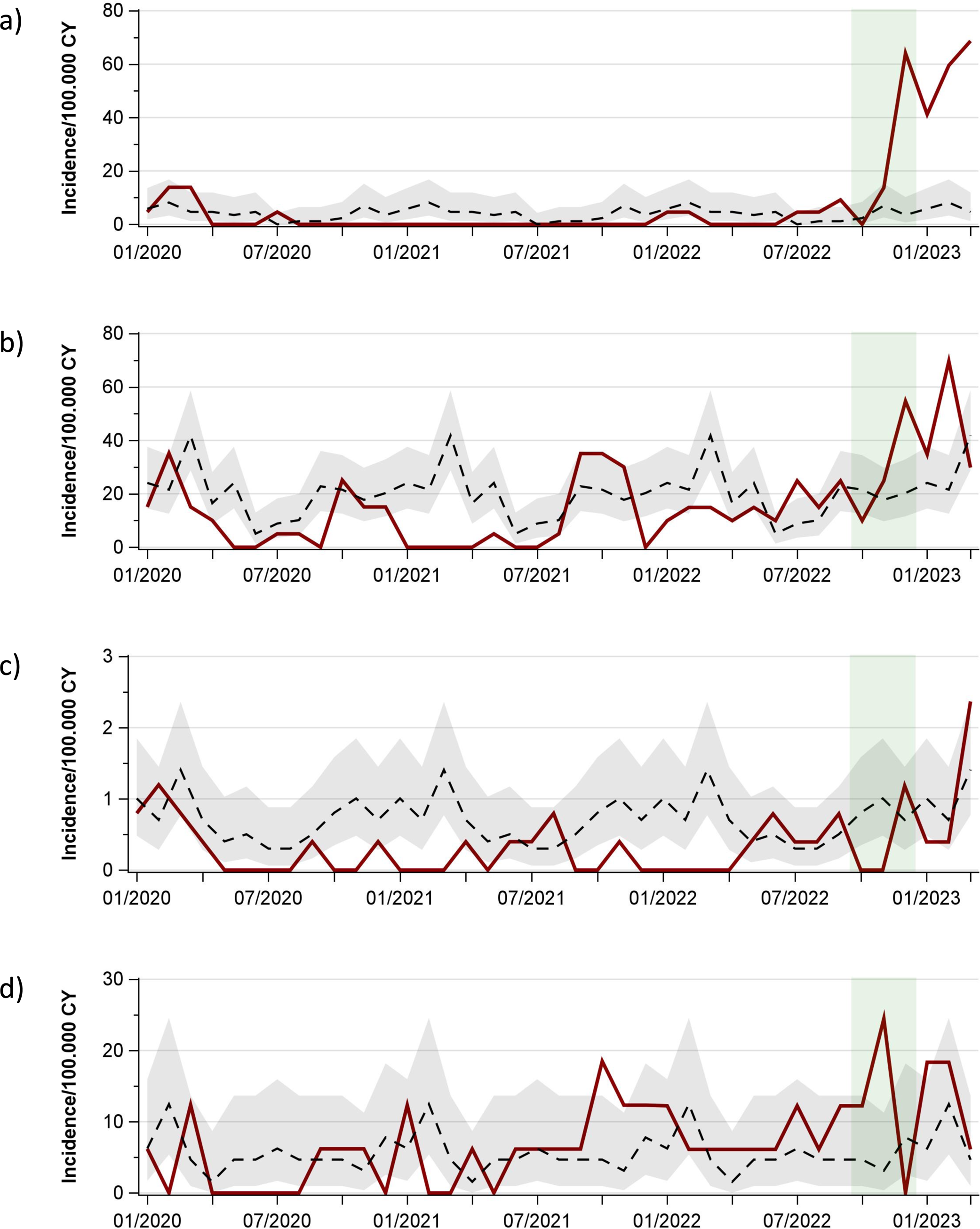
Hospitalizations for community-acquired bacterial infections. a) S. pyogenes b) S. pneumoniae c) N. meningitidis d) H. influenzae Red solid line: incidence rates January 2020 - March 2023; black dashed line and grey band: average monthly incidence rates 2016 - 2019 with 95 % CI; green band: period of MC study

**Figure 3.**
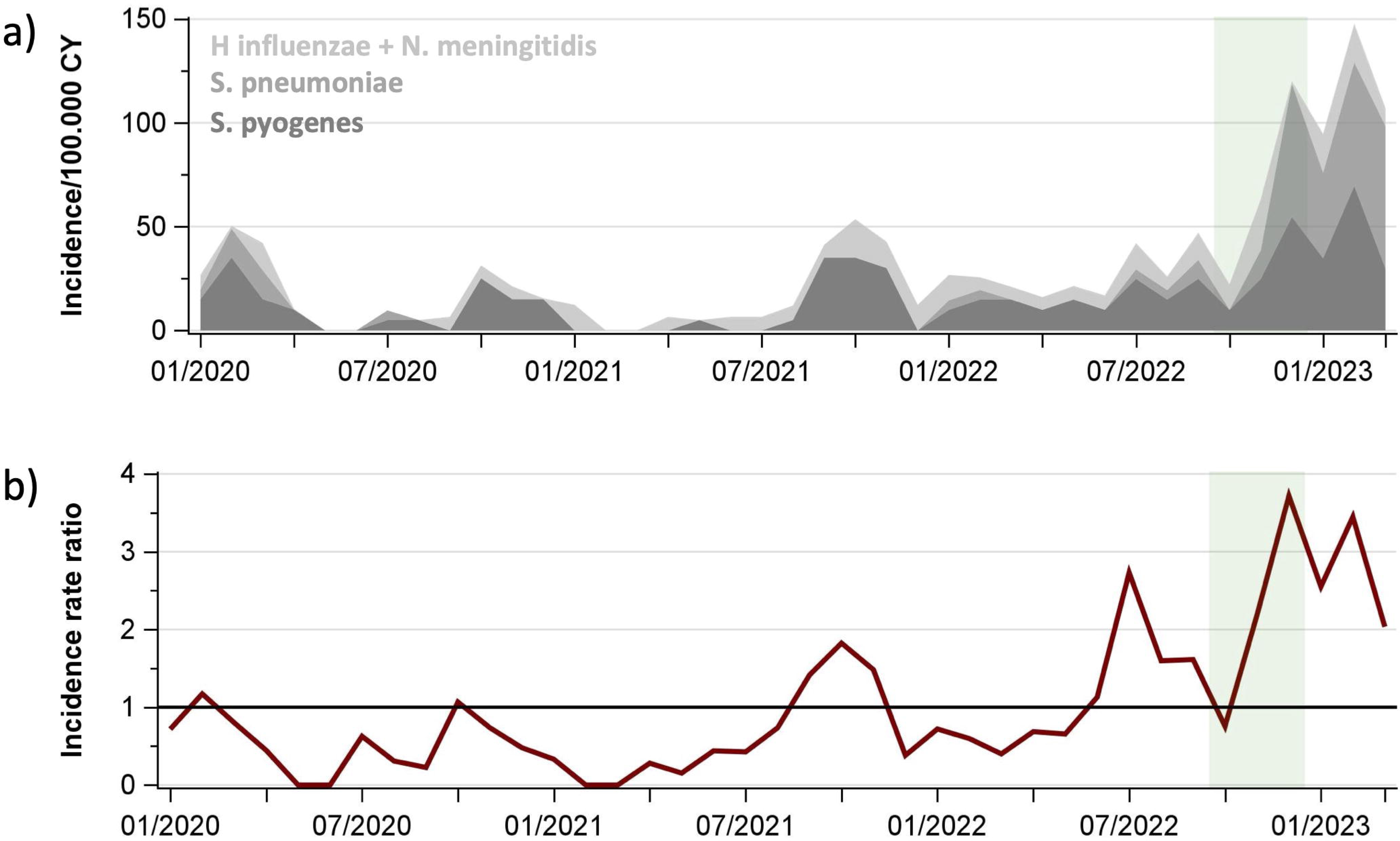
Hospitalizations due to acquired-bacterial infections from S. pyogenes, S. pneumoniae, N. meningitidis, and H. influenzae, January 2020 - March 2023. a) Cumulative hospitalization rates of S. pneumoniae (dark grey), S. pyogenes (medium grey), and N. meningitidis + H. influenzae (light grey) b) IRRs of infections, reference period 2016 – 2019 Green band: period of MC study

During the three-month MC study, the reported number of deaths in children < 15 years of age exceeded the expected number of deaths from bacterial infections for the entire year in NRW (7 observed; 1 expected). The causative pathogens were identified as *S. pyogenes* and *S. pneumoniae*, while no deaths were caused by *N. meningitidis* (2 expected) (Table 4).

## Discussion

This multi-source study in Germany’s most populous federal state detected a high burden of pediatric general ward and pediatric intensive care unit admissions for bacterial infections associated with *S. pyogenes, S. pneumoniae*, *N. meningitidis*, and *H. influenzae* in the last quarter of 2022. Disease severity was high with more than half of the cases admitted to an intensive care unit and frequent requirement of invasive mechanical ventilation and vasoactive agents. Longitudinal assessment since 2016 revealed an unprecedented peak of bacterial infection-related hospitalizations in late 2022 and early 2023, primarily driven by *S. pyogenes* and *S. pneumoniae*. Deaths during the three-month period of the MC study were seven times higher than expected for the entire year according to pre-pandemic death cause statistics.

A major finding that has already been reported in adults and children^13, 14^ was the marked overall decline of severe and invasive bacterial infections at the beginning of the COVID-19 pandemic. According to our study, this overall decline was shortly interrupted in fall 2021 but otherwise lasted until early summer 2022 when compulsory wearing of protective face masks was abrogated. However, decline patterns differed between the four bacterial pathogens examined in this study and were likely influenced by other factors than protective face masks.^13, 15–20^

The decline of *S. pyogenes* infections from the onset of the pandemic until spring 2022 was also reported in France, followed by a sudden increase of non-invasive *S. pyogenes* infections.^21^ Similar to Germany, France and England also witnessed strong increases of severe and invasive *S. pyogenes* infections during fall and winter of 2022.^7, 22^ These surges were most likely induced by an overall rise in infection numbers with the proportion of severe cases remaining constant, rather than changes in types, virulence or toxicity.^7, 23–25^ A possible explanation is the discontinuation of face mask wearing among children, which re-enabled droplet transmission of *S. pyogenes*, an essential mechanism of infection.^26^ Acute viral infections paving the way for bacterial superinfections are an additional factor potentially contributing to the severity of the *S. pyogenes* infection wave. This constellation is described for influenza virus in combination with *S. pyogenes* and *S. pneumoniae* ^27^ and was also observed in our study. The theory of superinfections as partial drivers of the surge is supported by the rise in consultations and hospitalizations for severe acute respiratory disease in children and adolescents during the peak of *S. pyogenes* infections in Germany in late 2022 reported by the Robert Koch-Institute.^28^

Viral infections likely also promoted the increase of *S. pneumoniae* infections. Asymptomatic *S. pneumoniae* carriage and respiratory virus infection as starting point for invasive pneumococcal disease is well-described.^29^ Our study found viral co-infections in 60 % of the *S. pneumoniae* cases, with influenza and respiratory syncytial virus as the most common. During the pandemic years, the decline in pneumococcal infections in Germany was less consistent compared to the drop in *S. pyogenes* infections, possibly explained by viral infection waves among children in spite of the pandemic measures.^28^ In contrast to *S. pyogenes* infections, invasive pneumococcal disease can be effectively prevented by vaccination.^30^ Consequently, it is crucial to establish and maintain high levels of vaccination rates to attenuate future outbreaks triggered by viral waves. In Germany, pneumococcal vaccination coverage for children entering primary school increased from 15 % in 2010 to 83 % in 2017-2019 but is as low as 2 % in the age group from 16 to 59 years, even in patients who suffer from high-risk clinical conditions.^31, 32^ Of infants born in 2016, only 73 % received a full series of the recommended pneumococcal vaccination scheme, vaccinations were frequently delayed, and nearly a quarter of infants did not receive a booster dose.^33^ This low vaccination coverage calls for urgent improvement to enhance protection against opportunistic pneumococcal infections during large-scale viral waves that are expected to persist in the post-SARS-CoV-2 era.

Vaccines are also responsible for decreasing incidence rates of meningococcal infections with marked replacement by non-vaccinated serogroups W and Y in several countries.^34^ In Germany, vaccination is recommended for serogroup C with an according decline of serogroup C infections since its introduction.^35^ The trend of overall decreasing meningococcal disease was further enhanced during the COVID-19 pandemic ^13^ and also noticeable in our study with incidence rates below pre-pandemic average. Yet, in March 2023, an increase in meningococcal infection incidence above average was observed but characterized by very low absolute numbers. In contrast to *S. pneumoniae*, vaccine coverage against meningococcal serogroup C in school starters in Germany is above 90 %.^32^ Unfortunately, no data are available on *N. meningitidis* serogroup B, but vaccine coverage is likely to be very low because vaccination is not officially recommended and thus not routinely covered by health insurances.

In the Netherlands, a decline of invasive infections during the COVID-19 pandemic was observed for *H. influenzae*, unfortunately associated with an increase of vaccine-preventable serotype b.^36^ In our study, *H. influenzae* hospitalizations during the pandemic remained within seasonally expected ranges, showing only two peaks in December of 2021 and 2022, respectively. However, despite vaccine coverage of 90 % in German children against *H. influenzae b* ^32^, IRRs have remained almost constantly above one since June 2021, indicating a slight increase of non-vaccinated *H. influenzae* infections that contribute to the overall burden of pediatric hospitalizations, especially during seasonal waves of acute respiratory diseases.

Limitations of the study mainly derive from missing cases: Due to the incomplete coverage of children’s hospitals in NRW, the MC study missed 6 % of pediatric hospital capacities. Cases treated outside of pediatric units, such as otorhinolaryngology or adult units, and pre-hospital deaths could not be captured, potentially leading to underestimation of incidences and disease severity. Variable pathogen-specific underreporting to the NRLs was also observed, which is acknowledged for infectious disease surveillance.^37–39^

In summary, the acute overload of the German pediatric health care system was caused by a post-pandemic rebound of acute viral and bacterial respiratory and invasive diseases that peaked at the same time, creating a fatal combination that caused an unprecedented incidence of hospitalizations in children. Shortages of essential drugs in the outpatient sector ^40, 41^ and a decrease in numbers of operable beds in children’s hospitals due to long-anticipated personnel shortages ^42, 43^ aggravated this pediatric health care crisis.

The results of this study present another piece of evidence that the side effects from preventive measures against the COVID-19 pandemic are at least on par with the harm children experienced from the novel coronavirus itself. While this is already well-acknowledged for mental health and obesity, which directly increase morbidity but not mortality ^44–47^, the wide-spread and drastic increase of severe and fatal bacterial infections in developed countries is new. Worldwide shortages of antibiotics for children ^40, 41^ complicated early treatment in the outpatient setting, which may have caused exacerbation of some cases. Despite good overall functional neurological outcomes observed in this study, non-neurological long-term sequelae, especially poststreptococcal autoimmune disorders, may continue to burden the health care system for years.

Further research should untangle this amalgam to quantify excess morbidity and mortality that result from acute bacterial infections themselves and from the failure to provide adequate pediatric care capacities in Germany. However, even more urgent than scientific workup, is the introduction of effective measures against future outbreaks among children to prevent morbidity and deaths. These include high vaccination coverage, opportunity to acquire natural immunity against viral and bacterial disease, sufficient pediatric drug supply, and pediatric health care capacities adapted to seasonal demands.

### Conclusion

The German pediatric health care crisis in late 2022 and early 2023 was caused by various mechanisms: an increased number of circulating bacterial pathogens, reduced adaptive immunity in the pediatric population following COVID-19 pandemic isolation measures, high burden of simultaneous viral infections, shortage of pediatric hospital capacities, and deficient pediatric antibiotic supply. Considering the upcoming infection season, an effective action plan including preventive measures needs to be combined with ongoing infection surveillance to prevent a possible new crisis in pediatric health care.

## Abbreviations

CI: confidence interval
CNS: central nervous system
CRC: capture-recapture
eCRF: web-based case report forms
ENT: ears nose and throat
FSO: Federal State Office *H. influenzae = Haemophilus influenzae*
ICD-10-GM: international classification of diseases 10 German modification
IQR: interquartile range
IRR: incidence rate ratios
MC: multicentre study, *N. meningitidis = Neisseria meningitidis,*
NRL: National Reference Laboratories
NRLMHi: National Reference Laboratory for *N. meningitidis* and *H. influenzae,*
NRW: North Rhine-Westphalia
PCPC: pediatric cerebral performance category
PICU: pediatric intensive care unit
rRNA: ribosomal Ribonucleic acid
RSV: respiratory syncytial virus, *S. pneumoniae = Streptococcus pneumoniae, S. pyrogenes = Streptococcus pyogenes,*
SIRS: systemic inflammatory response syndrome

## Article Summary

In late 2022 and early 2023, hospitalizations for bacterial infections dramatically increased, causing deaths and contributing to a near-collapse of the pediatric health care system.

## What’s Known on This Subject

During the COVID-19 pandemic, incidences of bacterial infections strongly declined. In late 2022, several European countries observed strongly rebounding incidences of invasive infections from group A streptococci.

## What This Study Adds

This study found up to 18-fold incidence rates of *S. pyogenes* in late 2022 and early 2023, accompanied by surges of *S. pneumoniae* and partially *H. influenzae*. Deaths were seven times higher than expected.

## Contributors Statement Page

Drs. Sarah C. Goretzki and Nora Bruns conceptualized and designed the study, carried out analyses, drafted the initial manuscript, and critically reviewed and revised the manuscript.

Drs Tom Hühne, Roland O. Adelmann, Firas Ala Eldin, Mohamed Albarouni, Jan-Claudius Becker, Michael A. Berghäuser, Thomas Boesing, Michael Boeswald, Milian Brasche, Francisco Brevis, Rokya Camara, Clara Deibert, Frank Dohle, Jörg Dolgner, Jan Dziobaka, Frank Eifinger, Natalie Elting, Matthias Endmann, Holger Frenzke, Monika Gappa, Bahman Gharavi, Christine Goletz, Eva Hahn, Yvonne Heidenreich, Konrad Heimann, Hans-Georg Hoffmann, Marc Hoppenz, Gerd Horneff, Helene Klassen, Cordula Körner-Rettberg, Pascal Lenz, Klaus Lohmeier, Andreas Müller, Frank Niemann, Michael Paulussen, Falk Pentek, Ruy Perez, Markus Pingel, Philip Repges, Tobias Rothoeft, Jochen Rübo, Herbert Schade, Robert Schmitz, Peter Schonhoff, Jan N. Schwade, Tobias Schwarz, Peter Seiffert, Georg Selzer, Uwe Spille, Carsten Thiel, Ansgar Thimm, Bartholomäus Urgatz, Alijda van den Heuvel, Tan van Hop, Verena Giesen, Stefan Wirth, Thomas Wollbrink, Daniel Wüller and Profs. Guido Engelmann, Kai O. Hensel, Alfred Längler, collected data and critically reviewed and revised the manuscript.

Drs. Mark van der Linden, Andreas Itzek, Heike Claus and Thiên-Trí Lâm collected data, carried analyses, and critically reviewed and revised the manuscript.

Profs. Ursula Felderhoff-Müser and Christian Dohna-Schwake conceptualized and designed the study, coordinated and supervised data collection, and critically reviewed and revised the manuscript for important intellectual content.

All authors approved the final manuscript as submitted and agree to be accountable for all aspects of the work.

## Data Availability

All data produced in the present study are available upon reasonable request to the authors.

## Acknowledgments

We thank Christina Pentek for her logistic support.

---

**Figure S1.**
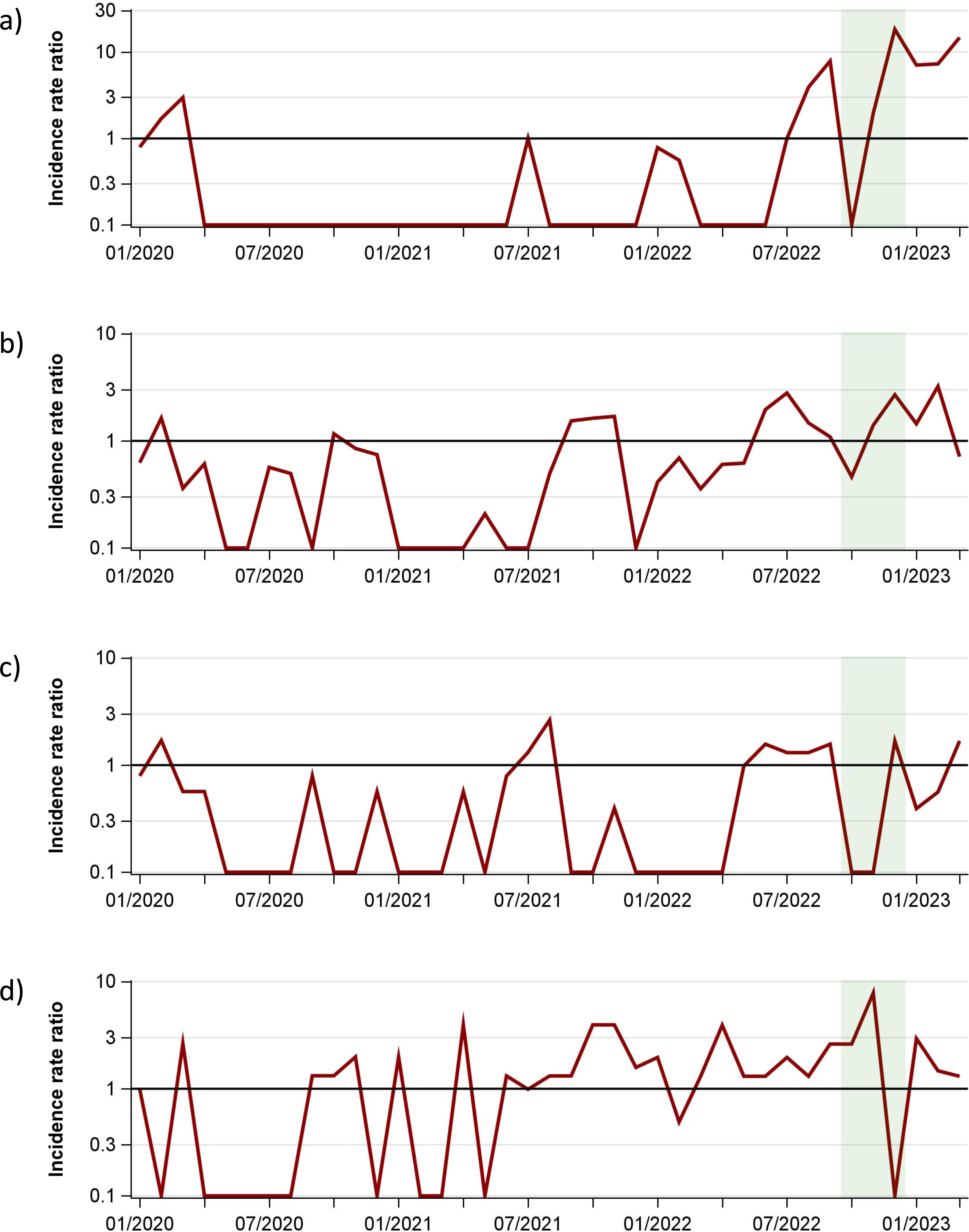
Monthly incidence rate ratios of invasive bacterial infections in children in North Rhine-Westphalia January 2020 -March 2023 (reference period 2016 – 2019) a) S. pyogenes b) S. pneumoniae c) N. meningitidis d) H. influenzae

