## Supplementary table for "Outbreak of severe community-acquired bacterial infections from *Streptococcus pyogenes, Streptococcus pneumoniae, Neisseria meningitidis*, and *Haemophilus influenzae* among children in North Rhine-Westphalia (Germany), October to December 2022"

| year | month | inc_StrepA | inc_StrepA_a | CI_lower_inc | CI_upper_inc | inc_Pneu |
|---|---|---|---|---|---|---|
| 2020 | 1 | 4,6 | 5,8 | 1,9 | 13,6 | 15,1 |
| 2020 | 2 | 13,9 | 8,2 | 3,3 | 16,8 | 35,2 |
| 2020 | 3 | 13,9 | 4,7 | 1,3 | 12,0 | 15,1 |
| 2020 | 4 | 0,0 | 4,7 | 1,3 | 12,0 | 10,1 |
| 2020 | 5 | 0,0 | 3,5 | 0,7 | 10,2 | 0,0 |
| 2020 | 6 | 0,0 | 4,7 | 1,3 | 12,0 | 0,0 |
| 2020 | 7 | 4,6 | 0,0 | 0,0 | 4,3 | 5,0 |
| 2020 | 8 | 0,0 | 1,2 | 0,0 | 6,5 | 5,0 |
| 2020 | 9 | 0,0 | 1,2 | 0,0 | 6,5 | 0,0 |
| 2020 | 10 | 0,0 | 2,3 | 0,3 | 8,4 | 25,1 |
| 2020 | 11 | 0,0 | 7,0 | 2,6 | 15,2 | 15,1 |
| 2020 | 12 | 0,0 | 3,5 | 0,7 | 10,2 | 15,1 |
| 2021 | 1 | 0,0 | 5,8 | 1,9 | 13,6 | 0,0 |
| 2021 | 2 | 0,0 | 8,2 | 3,3 | 16,8 | 0,0 |
| 2021 | 3 | 0,0 | 4,7 | 1,3 | 12,0 | 0,0 |
| 2021 | 4 | 0,0 | 4,7 | 1,3 | 12,0 | 0,0 |
| 2021 | 5 | 0,0 | 3,5 | 0,7 | 10,2 | 5,0 |
| 2021 | 6 | 0,0 | 4,7 | 1,3 | 12,0 | 0,0 |
| 2021 | 7 | 0,0 | 0,0 | 0,0 | 4,3 | 0,0 |
| 2021 | 8 | 0,0 | 1,2 | 0,0 | 6,5 | 5,0 |
| 2021 | 9 | 0,0 | 1,2 | 0,0 | 6,5 | 35,1 |
| 2021 | 10 | 0,0 | 2,3 | 0,3 | 8,4 | 35,1 |
| 2021 | 11 | 0,0 | 7,0 | 2,6 | 15,2 | 30,1 |
| 2021 | 12 | 0,0 | 3,5 | 0,7 | 10,2 | 0,0 |
| 2022 | 1 | 4,6 | 5,8 | 1,9 | 13,6 | 9,9 |
| 2022 | 2 | 4,6 | 8,2 | 3,3 | 16,8 | 14,9 |
| 2022 | 3 | 0,0 | 4,7 | 1,3 | 12,0 | 14,9 |
| 2022 | 4 | 0,0 | 4,7 | 1,3 | 12,0 | 9,9 |
| 2022 | 5 | 0,0 | 3,5 | 0,7 | 10,2 | 14,9 |
| 2022 | 6 | 0,0 | 4,7 | 1,3 | 12,0 | 9,9 |
| 2022 | 7 | 4,6 | 0,0 | 0,0 | 4,3 | 24,9 |
| 2022 | 8 | 4,6 | 1,2 | 0,0 | 6,5 | 14,9 |
| 2022 | 9 | 9,2 | 1,2 | 0,0 | 6,5 | 24,9 |
| 2022 | 10 | 0,0 | 2,3 | 0,3 | 8,4 | 9,9 |
| 2022 | 11 | 13,7 | 7,0 | 2,6 | 15,2 | 24,9 |
| 2022 | 12 | 64,1 | 3,5 | 0,7 | 10,2 | 54,7 |
| 2023 | 1 | 41,2 | 5,8 | 1,9 | 13,6 | 34,8 |
| 2023 | 2 | 59,5 | 8,2 | 3,3 | 16,8 | 69,6 |
| 2023 | 3 | 68,7 | 4,7 | 1,3 | 12,0 | 29,8 |

| inc_Pneu_av | CI_lower_inc | CI_upper_inc | inc_Men | inc_Men_ave | CI_lower_inc | CI_upper_inc |
|---|---|---|---|---|---|---|
| 24,1 | 14,5 | 37,6 | 0,8 | 1,0 | 0,5 | 1,8 |
| 21,5 | 12,6 | 34,5 | 1,2 | 0,7 | 0,3 | 1,5 |
| 41,8 | 28,8 | 58,7 | 0,8 | 1,4 | 0,8 | 2,4 |
| 16,5 | 8,8 | 28,2 | 0,4 | 0,7 | 0,3 | 1,5 |
| 24,1 | 14,5 | 37,6 | 0,0 | 0,4 | 0,1 | 1,0 |
| 5,1 | 1,4 | 13,0 | 0,0 | 0,5 | 0,2 | 1,2 |
| 8,9 | 3,6 | 18,3 | 0,0 | 0,3 | 0,1 | 0,9 |
| 10,1 | 4,4 | 20,0 | 0,0 | 0,3 | 0,1 | 0,9 |
| 22,8 | 13,5 | 36,1 | 0,4 | 0,5 | 0,2 | 1,2 |
| 21,5 | 12,6 | 34,5 | 0,0 | 0,8 | 0,3 | 1,6 |
| 17,7 | 9,7 | 29,8 | 0,0 | 1,0 | 0,5 | 1,8 |
| 20,3 | 11,6 | 32,9 | 0,4 | 0,7 | 0,3 | 1,5 |
| 24,1 | 14,5 | 37,6 | 0,0 | 1,0 | 0,5 | 1,8 |
| 21,5 | 12,6 | 34,5 | 0,0 | 0,7 | 0,3 | 1,5 |
| 41,8 | 28,8 | 58,7 | 0,0 | 1,4 | 0,8 | 2,4 |
| 16,5 | 8,8 | 28,2 | 0,4 | 0,7 | 0,3 | 1,5 |
| 24,1 | 14,5 | 37,6 | 0,0 | 0,4 | 0,1 | 1,0 |
| 5,1 | 1,4 | 13,0 | 0,4 | 0,5 | 0,2 | 1,2 |
| 8,9 | 3,6 | 18,3 | 0,4 | 0,3 | 0,1 | 0,9 |
| 10,1 | 4,4 | 20,0 | 0,8 | 0,3 | 0,1 | 0,9 |
| 22,8 | 13,5 | 36,1 | 0,0 | 0,5 | 0,2 | 1,2 |
| 21,5 | 12,6 | 34,5 | 0,0 | 0,8 | 0,3 | 1,6 |
| 17,7 | 9,7 | 29,8 | 0,4 | 1,0 | 0,5 | 1,8 |
| 20,3 | 11,6 | 32,9 | 0,0 | 0,7 | 0,3 | 1,5 |
| 24,1 | 14,5 | 37,6 | 0,0 | 1,0 | 0,5 | 1,8 |
| 21,5 | 12,6 | 34,5 | 0,0 | 0,7 | 0,3 | 1,5 |
| 41,8 | 28,8 | 58,7 | 0,0 | 1,4 | 0,8 | 2,4 |
| 16,5 | 8,8 | 28,2 | 0,0 | 0,7 | 0,3 | 1,5 |
| 24,1 | 14,5 | 37,6 | 0,4 | 0,4 | 0,1 | 1,0 |
| 5,1 | 1,4 | 13,0 | 0,8 | 0,5 | 0,2 | 1,2 |
| 8,9 | 3,6 | 18,3 | 0,4 | 0,3 | 0,1 | 0,9 |
| 10,1 | 4,4 | 20,0 | 0,4 | 0,3 | 0,1 | 0,9 |
| 22,8 | 13,5 | 36,1 | 0,8 | 0,5 | 0,2 | 1,2 |
| 21,5 | 12,6 | 34,5 | 0,0 | 0,8 | 0,3 | 1,6 |
| 17,7 | 9,7 | 29,8 | 0,0 | 1,0 | 0,5 | 1,8 |
| 20,3 | 11,6 | 32,9 | 1,2 | 0,7 | 0,3 | 1,5 |
| 24,1 | 14,5 | 37,6 | 0,4 | 1,0 | 0,5 | 1,8 |
| 21,5 | 12,6 | 34,5 | 0,4 | 0,7 | 0,3 | 1,5 |
| 41,8 | 28,8 | 58,7 | 2,4 | 1,4 | 0,8 | 2,4 |

| inc_HiB | inc_HiB_aver | CI_lower_inc | CI_upper_inc | IRR_StrepA | CI_upper_IRR | CI_lower_IRR |
|---|---|---|---|---|---|---|
| 6,2 | 6,2 | 1,7 | 16,0 | 0,8 | 2,4 | 0,3 |
| 0,0 | 12,5 | 5,4 | 24,6 | 1,7 | 4,2 | 0,8 |
| 12,4 | 4,7 | 1,0 | 13,7 | 3,0 | 10,9 | 1,2 |
| 0,0 | 1,6 | 0,0 | 8,7 | 0,0 | 0,0 | 0,0 |
| 0,0 | 4,7 | 1,0 | 13,7 | 0,0 | 0,0 | 0,0 |
| 0,0 | 4,7 | 1,0 | 13,7 | 0,0 | 0,0 | 0,0 |
| 0,0 | 6,2 | 1,7 | 16,0 | | | 1,1 |
| 0,0 | 4,7 | 1,0 | 13,7 | 0,0 | 0,0 | 0,0 |
| 6,2 | 4,7 | 1,0 | 13,7 | 0,0 | 0,0 | 0,0 |
| 6,2 | 4,7 | 1,0 | 13,7 | 0,0 | 0,0 | 0,0 |
| 6,2 | 3,1 | 0,4 | 11,3 | 0,0 | 0,0 | 0,0 |
| 0,0 | 7,8 | 2,5 | 18,2 | 0,0 | 0,0 | 0,0 |
| 12,3 | 6,2 | 1,7 | 16,0 | 0,0 | 0,0 | 0,0 |
| 0,0 | 12,5 | 5,4 | 24,6 | 0,0 | 0,0 | 0,0 |
| 0,0 | 4,7 | 1,0 | 13,7 | 0,0 | 0,0 | 0,0 |
| 6,2 | 1,6 | 0,0 | 8,7 | 0,0 | 0,0 | 0,0 |
| 0,0 | 4,7 | 1,0 | 13,7 | 0,0 | 0,0 | 0,0 |
| 6,2 | 4,7 | 1,0 | 13,7 | 0,0 | 0,0 | 0,0 |
| 6,2 | 6,2 | 1,7 | 16,0 | | | 0,0 |
| 6,2 | 4,7 | 1,0 | 13,7 | 0,0 | 0,0 | 0,0 |
| 6,2 | 4,7 | 1,0 | 13,7 | 0,0 | 0,0 | 0,0 |
| 18,5 | 4,7 | 1,0 | 13,7 | 0,0 | 0,0 | 0,0 |
| 12,3 | 3,1 | 0,4 | 11,3 | 0,0 | 0,0 | 0,0 |
| 12,3 | 7,8 | 2,5 | 18,2 | 0,0 | 0,0 | 0,0 |
| 12,2 | 6,2 | 1,7 | 16,0 | 0,8 | 2,4 | 0,3 |
| 6,1 | 12,5 | 5,4 | 24,6 | 0,6 | 1,4 | 0,3 |
| 6,1 | 4,7 | 1,0 | 13,7 | 0,0 | 0,0 | 0,0 |
| 6,1 | 1,6 | 0,0 | 8,7 | 0,0 | 0,0 | 0,0 |
| 6,1 | 4,7 | 1,0 | 13,7 | 0,0 | 0,0 | 0,0 |
| 6,1 | 4,7 | 1,0 | 13,7 | 0,0 | 0,0 | 0,0 |
| 12,2 | 6,2 | 1,7 | 16,0 | | | 1,1 |
| 6,1 | 4,7 | 1,0 | 13,7 | 3,9 | 154,9 | 0,7 |
| 12,2 | 4,7 | 1,0 | 13,7 | 7,8 | 309,9 | 1,4 |
| 12,2 | 4,7 | 1,0 | 13,7 | 0,0 | 0,0 | 0,0 |
| 24,5 | 3,1 | 0,4 | 11,3 | 2,0 | 5,3 | 0,9 |
| 0,0 | 7,8 | 2,5 | 18,2 | 18,3 | 88,8 | 6,3 |
| 18,4 | 6,2 | 1,7 | 16,0 | 7,1 | 21,7 | 3,0 |
| 18,4 | 12,5 | 5,4 | 24,6 | 7,3 | 18,1 | 3,5 |
| 6,1 | 4,7 | 1,0 | 13,7 | 14,7 | 54,0 | 5,7 |

| IRR_Pneu | CI_upper_IRF | CI_lower_IRF | IRR_Men | CI_upper_IRF | CI_lower_IRF | IRR_HiB |
|---|---|---|---|---|---|---|
| 0,6 | 1,0 | 0,4 | 0,8 | 1,7 | 0,4 | 1,0 |
| 1,6 | 2,8 | 1,0 | 1,7 | 4,2 | 0,8 | 0,0 |
| 0,4 | 0,5 | 0,3 | 0,6 | 1,0 | 0,3 | 2,6 |
| 0,6 | 1,1 | 0,4 | 0,6 | 1,4 | 0,3 | 0,0 |
| 0,0 | 0,0 | 0,0 | 0,0 | 0,0 | 0,0 | 0,0 |
| 0,0 | 0,0 | 0,0 | 0,0 | 0,0 | 0,0 | 0,0 |
| 0,6 | 1,4 | 0,3 | 0,0 | 0,0 | 0,0 | 0,0 |
| 0,5 | 1,1 | 0,3 | 0,0 | 0,0 | 0,0 | 0,0 |
| 0,0 | 0,0 | 0,0 | 0,8 | 2,4 | 0,3 | 1,3 |
| 1,2 | 2,0 | 0,7 | 0,0 | 0,0 | 0,0 | 1,3 |
| 0,8 | 1,6 | 0,5 | 0,0 | 0,0 | 0,0 | 2,0 |
| 0,7 | 1,3 | 0,5 | 0,6 | 1,4 | 0,3 | 0,0 |
| 0,0 | 0,0 | 0,0 | 0,0 | 0,0 | 0,0 | 2,0 |
| 0,0 | 0,0 | 0,0 | 0,0 | 0,0 | 0,0 | 0,0 |
| 0,0 | 0,0 | 0,0 | 0,0 | 0,0 | 0,0 | 0,0 |
| 0,0 | 0,0 | 0,0 | 0,6 | 1,4 | 0,3 | 4,0 |
| 0,2 | 0,3 | 0,1 | 0,0 | 0,0 | 0,0 | 0,0 |
| 0,0 | 0,0 | 0,0 | 0,8 | 2,4 | 0,3 | 1,3 |
| 0,0 | 0,0 | 0,0 | 1,3 | 6,4 | 0,5 | 1,0 |
| 0,5 | 1,1 | 0,3 | 2,6 | 12,8 | 0,9 | 1,3 |
| 1,5 | 2,6 | 1,0 | 0,0 | 0,0 | 0,0 | 1,3 |
| 1,6 | 2,8 | 1,0 | 0,0 | 0,0 | 0,0 | 4,0 |
| 1,7 | 3,1 | 1,0 | 0,4 | 0,8 | 0,2 | 4,0 |
| 0,0 | 0,0 | 0,0 | 0,0 | 0,0 | 0,0 | 1,6 |
| 0,4 | 0,7 | 0,3 | 0,0 | 0,0 | 0,0 | 2,0 |
| 0,7 | 1,2 | 0,4 | 0,0 | 0,0 | 0,0 | 0,5 |
| 0,4 | 0,5 | 0,3 | 0,0 | 0,0 | 0,0 | 1,3 |
| 0,6 | 1,1 | 0,4 | 0,0 | 0,0 | 0,0 | 3,9 |
| 0,6 | 1,0 | 0,4 | 1,0 | 3,6 | 0,4 | 1,3 |
| 2,0 | 7,2 | 0,8 | 1,6 | 4,8 | 0,7 | 1,3 |
| 2,8 | 7,0 | 1,4 | 1,3 | 6,3 | 0,4 | 2,0 |
| 1,5 | 3,4 | 0,7 | 1,3 | 6,3 | 0,4 | 1,3 |
| 1,1 | 1,8 | 0,7 | 1,6 | 4,8 | 0,7 | 2,6 |
| 0,5 | 0,8 | 0,3 | 0,0 | 0,0 | 0,0 | 2,6 |
| 1,4 | 2,6 | 0,8 | 0,0 | 0,0 | 0,0 | 7,8 |
| 2,7 | 4,7 | 1,7 | 1,7 | 4,2 | 0,8 | 0,0 |
| 1,4 | 2,4 | 0,9 | 0,4 | 0,8 | 0,2 | 2,9 |
| 3,2 | 5,5 | 2,0 | 0,6 | 1,4 | 0,3 | 1,5 |
| 0,7 | 1,0 | 0,5 | 1,7 | 3,1 | 1,0 | 1,3 |

| CI_upper_IRR_ | CI_lower_IRR | incidence_rat | incidence_cur | IRR_cum |
|---|---|---|---|---|
| 3,6 | 0,4 | 26,7 | 37,2 | 0,7 |
| 0,0 | 0,0 | 50,3 | 42,9 | 1,2 |
| 12,8 | 0,9 | 42,1 | 52,6 | 0,8 |
| 0,0 | 0,0 | 10,5 | 23,4 | 0,4 |
| 0,0 | 0,0 | 0,0 | 32,7 | 0,0 |
| 0,0 | 0,0 | 0,0 | 14,9 | 0,0 |
| 0,0 | 0,0 | 9,7 | 15,4 | 0,6 |
| 0,0 | 0,0 | 5,0 | 16,3 | 0,3 |
| 6,4 | 0,5 | 6,6 | 29,2 | 0,2 |
| 6,4 | 0,5 | 31,3 | 29,4 | 1,1 |
| 16,4 | 0,5 | 21,3 | 28,9 | 0,7 |
| 0,0 | 0,0 | 15,5 | 32,3 | 0,5 |
| 7,3 | 0,8 | 12,3 | 37,2 | 0,3 |
| 0,0 | 0,0 | 0,0 | 42,9 | 0,0 |
| 0,0 | 0,0 | 0,0 | 52,6 | 0,0 |
| 156,1 | 0,7 | 6,6 | 23,4 | 0,3 |
| 0,0 | 0,0 | 5,0 | 32,7 | 0,2 |
| 6,4 | 0,5 | 6,6 | 14,9 | 0,4 |
| 3,6 | 0,4 | 6,6 | 15,4 | 0,4 |
| 6,4 | 0,5 | 12,0 | 16,3 | 0,7 |
| 6,4 | 0,5 | 41,2 | 29,2 | 1,4 |
| 19,2 | 1,4 | 53,6 | 29,4 | 1,8 |
| 32,6 | 1,1 | 42,8 | 28,9 | 1,5 |
| 4,9 | 0,7 | 12,3 | 32,3 | 0,4 |
| 7,2 | 0,8 | 26,8 | 37,2 | 0,7 |
| 1,1 | 0,2 | 25,6 | 42,9 | 0,6 |
| 6,3 | 0,4 | 21,0 | 52,6 | 0,4 |
| 154,9 | 0,7 | 16,1 | 23,4 | 0,7 |
| 6,3 | 0,4 | 21,4 | 32,7 | 0,7 |
| 6,3 | 0,4 | 16,9 | 14,9 | 1,1 |
| 7,2 | 0,8 | 42,1 | 15,4 | 2,7 |
| 6,3 | 0,4 | 26,0 | 16,3 | 1,6 |
| 12,7 | 0,9 | 47,0 | 29,2 | 1,6 |
| 12,7 | 0,9 | 22,2 | 29,4 | 0,8 |
| 64,8 | 2,2 | 63,1 | 28,9 | 2,2 |
| 0,0 | 0,0 | 120,0 | 32,3 | 3,7 |
| 10,8 | 1,1 | 94,8 | 37,2 | 2,6 |
| 3,4 | 0,7 | 147,9 | 42,9 | 3,4 |
| 6,3 | 0,4 | 107,0 | 52,6 | 2,0 |

| | |
|---|---|
| **inc_StrepA** | incidence rate of S. pyogenes per 100.000 child years |
| **inc_StrepA_average_2016_2019** | average incidence rate of S. pyogenes per 100.000 child y |
| **CI_lower_inc_StrepA_average_2016_201** | lower confidence limit of average incidence rate of S. pyo |
| **CI_upper_inc_StrepA_average_2016_201** | upper confidence limit of average incidence rate of S. pyo |
| **inc_Pneu** | incidence rate of S. pneumoniae per 100.000 child years |
| **inc_Pneu_average_2016_2019** | average incidence rate of S. pneumoniae per 100.000 chil |
| **CI_lower_inc_Pneu_average_2016_2019** | lower confidence limit of average incidence rate of S. pne |
| **CI_upper_inc_Pneu_average_2016_2019** | upper confidence limit of average incidence rate of S. pne |
| **inc_Men** | incidence rate of N. meningitidis per 100.000 child years |
| **inc_Men_average_2016_2019** | average incidence rate of N. meningitidis per 100.000 chil |
| **CI_lower_inc_Men_average_2016_2019** | lower confidence limit of average incidence rate of N. me |
| **CI_upper_inc_Men_average_2016_2019** | upper confidence limit of average incidence rate of N. me |
| **inc_HiB** | incidence rate of H. influenzae per 100.000 child years |
| **inc_HiB_average_2016_2019** | average incidence rate of H. influenzae per 100.000 child |
| **CI_lower_inc_HiB_average_2016_2019** | lower confidence limit of average incidence rate of H. infl |
| **CI_upper_inc_HiB_average_2016_2019** | upper confidence limit of average incidence rate of H. infl |
| **IRR_StrepA** | monthly incidence rate ratio of S. pyogenes (reference pe |
| **CI_upper_IRR_StrepA** | upper confidence limit of IRR of S. pyogenes |
| **CI_lower_IRR_StrepA** | lower confidence limit of IRR of S. pyogenes |
| **IRR_Pneu** | monthly incidence rate ratio of S. pneumoniae (reference |
| **CI_upper_IRR_Pneu** | upper confidence limit of IRR of S. pneumoniae |
| **CI_lower_IRR_Pneu** | lower confidence limit of IRR of S. pneumoniae |
| **IRR_Men** | monthly incidence rate ratio of N. meningitidis (reference |
| **CI_upper_IRR_Men** | upper confidence limit of IRR of N. meningitidis |
| **CI_lower_IRR_Men** | lower confidence limit of IRR of N. meningitidis |
| **IRR_HiB** | monthly incidence rate ratio of H. influenzae (reference p |
| **CI_upper_IRR_HiB** | upper confidence limit of IRR of H. influenzae |
| **CI_lower_IRR_HiB** | lower confidence limit of IRR of H. influenzae |

genes per 100.000 child years from 2016 - 2019
genes per 100.000 child years from 2016 - 2019

umoniae per 100.000 child years from 2016 - 2019
umoniae per 100.000 child years from 2016 - 2019

ningitidis per 100.000 child years from 2016 - 2019
ningitidis per 100.000 child years from 2016 - 2019

uenzae per 100.000 child years from 2016 - 2019
luenzae per 100.000 child years from 2016 - 2019
